# Dynamics of Influenza A and SARS-CoV-2 coinfections during the COVID-19 pandemic in India

**DOI:** 10.1101/2023.02.19.23285730

**Authors:** Sandhra Ravikumar, Ekant Tamboli, Shefali Rahangdale, Lekha Salsekar, Siddharth Singh Tomar, Krishna Khairnar

**Author notes:** Corresponding author: Correspondence to Krishna Khairnar.

## Abstract

The SARS-CoV-2 exhibits similar aetiology, mode of transmission and clinical presentation as the H1N1pdm09 (a subtype of Influenza A) and Influenza A (other subtypes), and can exist as a coinfection in the same patient. It is essential to understand the coinfection dynamics of these viruses for effective management of the disease. This study examined 959 SARS-CoV-2 positive samples collected from the six states and three union territories in India from May to December 2022. The clinical data was accessed from the Integrated Health Information Platform (IHIP) and the Indian council of medical research (ICMR) COVID-19 data portal. The samples were tested for SARS-CoV-2, H1N1pdm09 and Influenza A using Reverse Transcriptase Real-Time Polymerase Chain Reaction q(RT-PCR). All 959 samples were subjected to SARS-CoV-2 whole genome sequencing (WGS) using Oxford Nanopore Next Generation Sequencing (NGS). From the 959 SARS-CoV-2 positive samples, 17.5% were co-infected with H1N1pdm09, 8.2% were co-infected with Influenza A, and 74.2% were only positive for SARS-CoV-2. The comparative analysis of viral load among the coinfected cases revealed that Influenza A and H1N1pdm09 had higher viral loads than SARS-CoV-2 in the studied samples. Out of 959 samples subjected to WGS, 815 and 144 were considered quality control (QC) passed, and QC failed, respectively, for SARS-CoV-2 variant calling. SARS-CoV-2 WGS identified 46 different variants belonging to the Omicron lineage. The SARS-CoV-2 and Influenza A coinfection group; and the SARS-CoV-2 and H1N1pdm09 coinfection group showed a higher proportion of symptomatic cases. This work demonstrates the need for coinfection analysis for the H1N1pdm09 virus, Influenza A virus and SARS-CoV-2 while studying the etiological agent in individuals with ILI/SARI symptoms. It is recommended that, in addition to determining the aetiology of ILI/SARI, an examination for H1N1pdm09 and Influenza A be conducted concurrently utilising molecular tools such as WGS and RT-PCR to understand the variant dynamics and the viral load for taking an informed decision during the patient management and treatment discourse.

## 1. INTRODUCTION

The severe acute respiratory syndrome coronavirus 2 (SARS-CoV-2) pandemic took the world by storm in December 2019. Soon it was declared a pandemic by the World Health Organization (WHO). The SARS-CoV-2 virus infects the respiratory system and causes influenza-like or severe acute respiratory illness (ILI/SARI) symptoms. However, SARS-CoV-2 is not the only pandemic faced by humans.

Instances of respiratory virus outbreaks attaining pandemic status have been frequent throughout history. Russian influenza that lasted from 1889 to 1892 [1], Spanish influenza of 1918–1920 [2], Asian influenza (H2N2) that appeared during 1957–1958 [3], the spells of SARS from 2002-2003 [4], the Swine flu (H1N1) pandemic of 2009 [5] and the middle east respiratory syndrome related-coronavirus (MERS-CoV) of 2012 [6] to name a few.

India has faced the deadly second wave of the SARS-CoV-2 pandemic and is currently overcoming the human and financial loss caused by it. It is also important to mention that from August to November 2022, while the SARS-CoV-2 pandemic was still ongoing, the H1N1 (a subtype of Influenza A of Swine-origin commonly known as the Swine flu) cases also started increasing in India, particularly, in the states of Gujarat, Maharashtra, and Tamil Nadu [7]. This overlapping of an outbreak and an ongoing pandemic encouraged us to investigate the co-prevalence/coinfections of respiratory viruses, such as Influenza A and H1N1 pandemic 2009 (H1N1pdm09), with SARS-CoV-2 viruses in the population. Influenza viruses belong to the family *Orthomyxoviridae* and have 4 subtypes; A (*Alphainfluenzavirus*), B (*Betainfluenzavirus*), C (*Gammainfluenzavirus*), and D *(Deltainfluenzavirus*). Influenza A, B, and C can infect humans, while Influenza D infects animals, particularly cattle [8]. Influenza B can infect Humans and Seals, but this virus is not known to cause pandemics [9]. Therefore, Influenza A remains an epidemiologically significant type of Influenza virus that can be classified into subtypes based on different types of hemagglutinin and neuraminidase surface proteins. H1N1 and H3N2 are widely circulating subtypes of Influenza A [8]. Influenza viruses have a similar mode of transmission and clinical presentation as the SARS-CoV-2 virus; therefore, the coinfection analysis is significant in resolving the aetiology [10]. A study by Kinoshita et. al. showed that coinfection could contribute to aggravating the symptoms and increasing the severity of the disease [11]. Therefore, it is essential to identify a coinfection and make an informed clinical decision to manage the disease. In patients where co-infection of SARS-CoV-2 with Influenza A and/or H1N1pdm09 is detected, a symptomatic manifestation of ILI could also be attributed to H1N1pdm09 or Influenza A as the patient is vaccinated against SARS-CoV-2, but not against Influenza. In the case of SARS-CoV-2 vaccine escape, the symptomatic manifestation in coinfection cases could either be due to the vaccine-escape strain of SARS-CoV-2 or the Influenza virus. A study is warranted to understand how the clinical presentations manifest differently in coinfection cases, it could also give some interesting insights into the symptomatic and asymptomatic distribution of cases among the given population.

Therefore, we attempted to investigate the dynamics of H1N1pdm09 or Influenza A coinfections in SARS-CoV-2 positive cases. As per the National Center for Disease Control (NCDC) data for 2022, the tally of H1N1pdm09 cases in India stands at 13,202; and 402 deaths are attributed to H1N1pdm09 infection. The states of Gujarat, Maharashtra, and Tamil Nadu were worst affected. Gujarat recorded 2174 cases and 71 deaths, Maharashtra had 3714 cases and 215 deaths, and Tamil Nadu had 2827 cases and 25 deaths [7]. The caseload in these states increased from August to November 2022 [12,13,14,15]. Therefore the nasopharyngeal and oropharyngeal swab samples from this period were investigated retrospectively for detecting SARS-CoV-2 with Influenza A and H1N1pdm09 coinfections.

## 2. MATERIALS AND METHODS

### 2.1 Data collection

The metadata of 959 SARS-CoV-2 positive patients from May to December 2022 was taken from the Integrated Health Information Platform (IHIP), a web portal made as a part of the Integrated Disease Surveillance Programme (IDSP), India. The IHIP web portal was established and maintained by the Ministry of Health and Family Welfare, Government of India. It is used by sentinel surveillance sites such as hospitals, virus research and diagnostics laboratories to record patient metadata during the SARS-CoV-2 genomic surveillance [16]. These SARS-CoV-2 positive samples were selected for Influenza A and H1N1pdm09 testing since the Influenza A and H1N1pdm09 caseload was high during this period in India. The metadata of SARS-CoV-2 positive cases included in this study includes patient details such as age, gender, quantitative reverse transcriptase real-time polymerase chain reaction (qRT-PCR) cycle threshold (Ct) value, and SARS-CoV-2 variant information. The symptomatic or asymptomatic status of the patients was sought from the Indian council of medical research (ICMR) COVID-19 data portal.

### 2.2 Materials

The RNA extraction was performed using Insta NX automated nucleic acid extractor. QuantStudio 3 and StepOnePlus qRT-PCR from Applied Biosystems machines were used for qRT-PCR, cDNA synthesis, PCR tiling, and rapid barcoding. The nucleic acid quantification for quality control (QC) of the DNA libraries for Oxford Nanopore Technology (ONT) based whole genome sequencing (WGS) was performed using a Qubit 4 Fluorometer from Thermo Fisher Scientific. SARS-CoV-2 WGS was performed using an ONT MinION sequencing platform of Mk1C 6.3.9 and Mk1B.

### 2.3 Sample collection and RNA extraction

The 959 SARS-CoV-2 positive samples collected from May to December 2022 were from 6 states and 3 Union territories of India, namely, Maharashtra, Tamil Nadu, Madhya Pradesh, Kerala, Haryana, and Chhattisgarh; and 3 Union territories of India, namely Jammu and Kashmir, Puducherry, and Delhi. The states of Maharashtra (n=792), Tamil Nadu (n=123), and Jammu & Kashmir (n=29) had a significant contribution of 98.4% among the total samples. Overall gender distribution in the sample set was 518 (54.02%) males and 441 (45.98%) females. In this study, the number of samples distributed within the age group was 1 (0 to <2 years), 6 (2 to <5 years), 34 (5 to <15 years), 616 (15 to <50 years), 202 (50 to <65 years), and 100 (≥ 65 years). The age distribution was according to the WHO Global Epidemiological Surveillance Standards for Influenza [17]. The sample collection was carried out using a nasopharyngeal-oropharyngeal (NP/OP) swab suspended in a viral transport medium (VTM). The RNA from VTM swab samples was extracted using an automated RNA extractor using MBIN013 Insta NX Viral RNA Purification Kit as per the manufacturer’s protocol (HIMEDIA) [18]. The extracted RNA samples were used immediately or stored at -80°C till further use.

### 2.4 SARS-CoV-2 whole genome sequencing

The SARS-CoV-2 positive samples with Ct values of ≤ 25 were selected for SARS-CoV-2 WGS. The cDNA synthesis was carried out using the TAKARA Prime Script RT reagent kit (RR037A) [19]. The sequencing libraries were constructed by multiplex PCR tiling according to the protocol of ONT [20]. The prepared cDNA libraries were sequenced using MinION Mk1C or MinION Mk1B.

### 2.5 Bioinformatic analysis

The live base-calling was performed using the Guppyv22.10.7 base-calling algorithm integrated into the MinION Mk1C sequencer [21]. The processed FASTQ reads from the MinION sequencer were analysed using the bioinformatics platform COMMANDER developed by Genotypic Technology Pvt. Ltd [22]. COMMANDER is a graphical user interface software developed to ease bioinformatic analysis post-sequencing. The FASTA sequence and the variant call performed by the COMMANDER pipeline were further confirmed by the web-based Pangolin COVID-19 Lineage Assigner [23]. The metadata, including the FASTA sequence, was submitted to the Global initiative on sharing Avian Influenza Data (GISAID), and the Indian biological data centre (IBDC).

### 2.6 RT-PCR for detecting SARS-CoV-2, Influenza A and H1N1pdm09

qRT-PCR was performed for quantitative detection of SARS-CoV-2 using MBPCR255 Hi-PCR COVID-19 Triplex Probe PCR Kit (HIMEDIA), and Influenza A and H1N1pdm09 using MBPCR263 Hi-PCR Influenza Multiplex Probe PCR kit (HIMEDIA) in each sample. The primer-probe sets in the kits were specific to detect viral RNA from SARS-CoV-2, Influenza A and H1N1pdm09. The samples with a Ct value of ≤ 38 were considered RT-PCR positive.

## 3. RESULTS

### 3.1 The coinfection of SARS-CoV-2 and Influenza A or H1N1pdm09

Out of 959 SARS-CoV-2 positive samples, 168 (17.5%) samples were found to be coinfected with H1N1pdm09, and 79 (8.2%) were found to be coinfected with Influenza A; whereas 712 (74.2%) samples were solely positive for SARS-CoV-2 (mono-infection) (**Fig.1**). While analysing the trend of the coinfected cases within the timeline of this study, it was observed that June had the highest number of coinfection cases followed by July and August. The number of coinfection cases was the least in May, October, September, December, and November. The SARS-CoV-2 and Influenza A coinfected cases were observed to be the highest in June, August, and May, while the SARS-CoV-2 and H1N1pdm09 coinfected cases were highest in June, July, and August.

**Figure 1:**
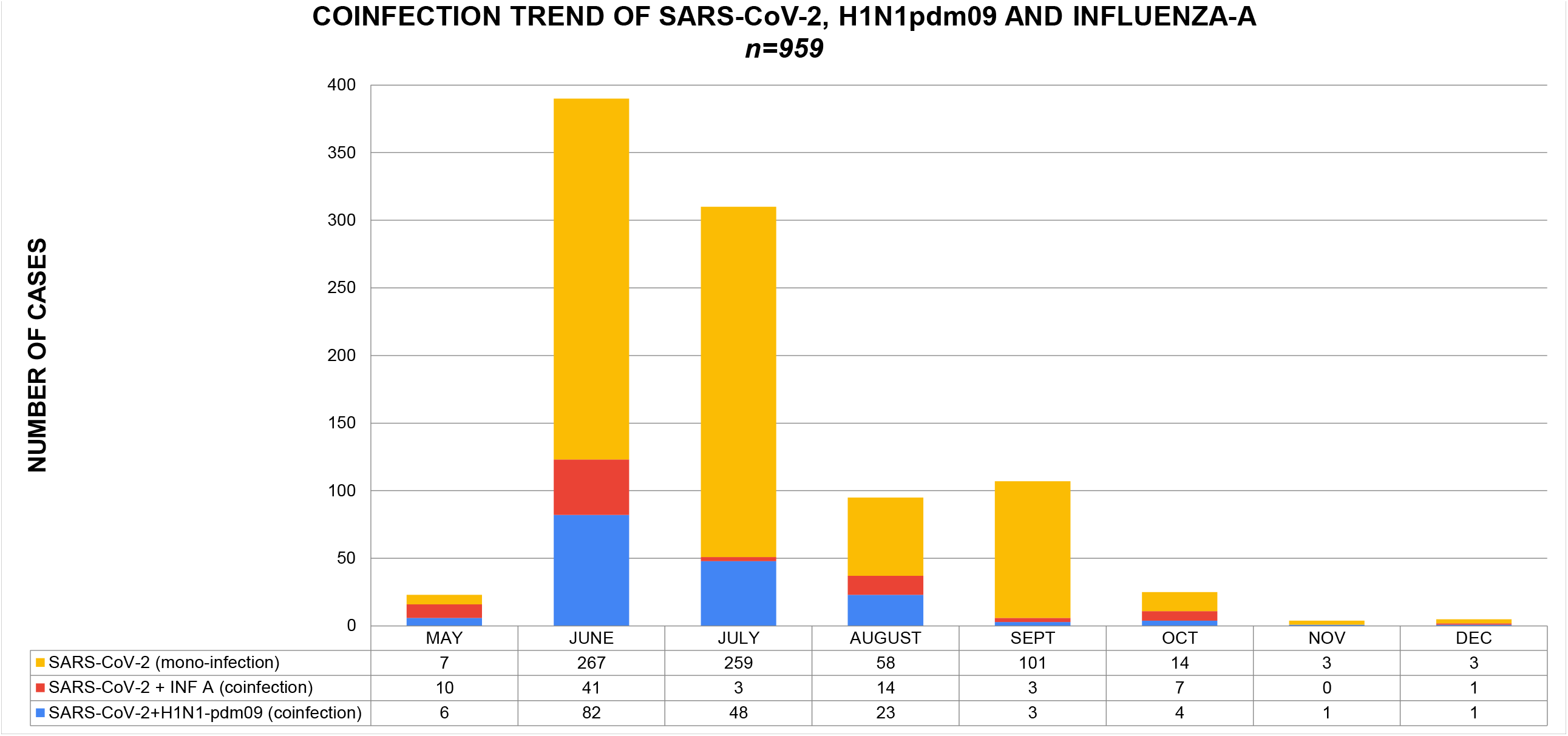
Month-wise prevalence trend of SARS-CoV-2 and H1N1pdm09 coinfection (Blue); SARS-CoV-2 and Influenza A coinfection (Red); and SARS-CoV-2 mono-infection (Yellow) in studied cases (n=959).

The highest number of coinfection cases was found to be in the age group of 15 to <50 years, followed by 50 to <65 years, ≥ 65 years, and 5 to <15 years (**Fig.2**). Among these age groups, it was also observed that the proportionality of SARS-CoV-2 coinfection with H1N1pdm09 was higher in comparison to SARS-CoV-2 coinfection with Influenza A. However, in the age groups of 2 to <5 years, the proportionality of coinfected cases was found to be relatively low.

**Figure 2:**
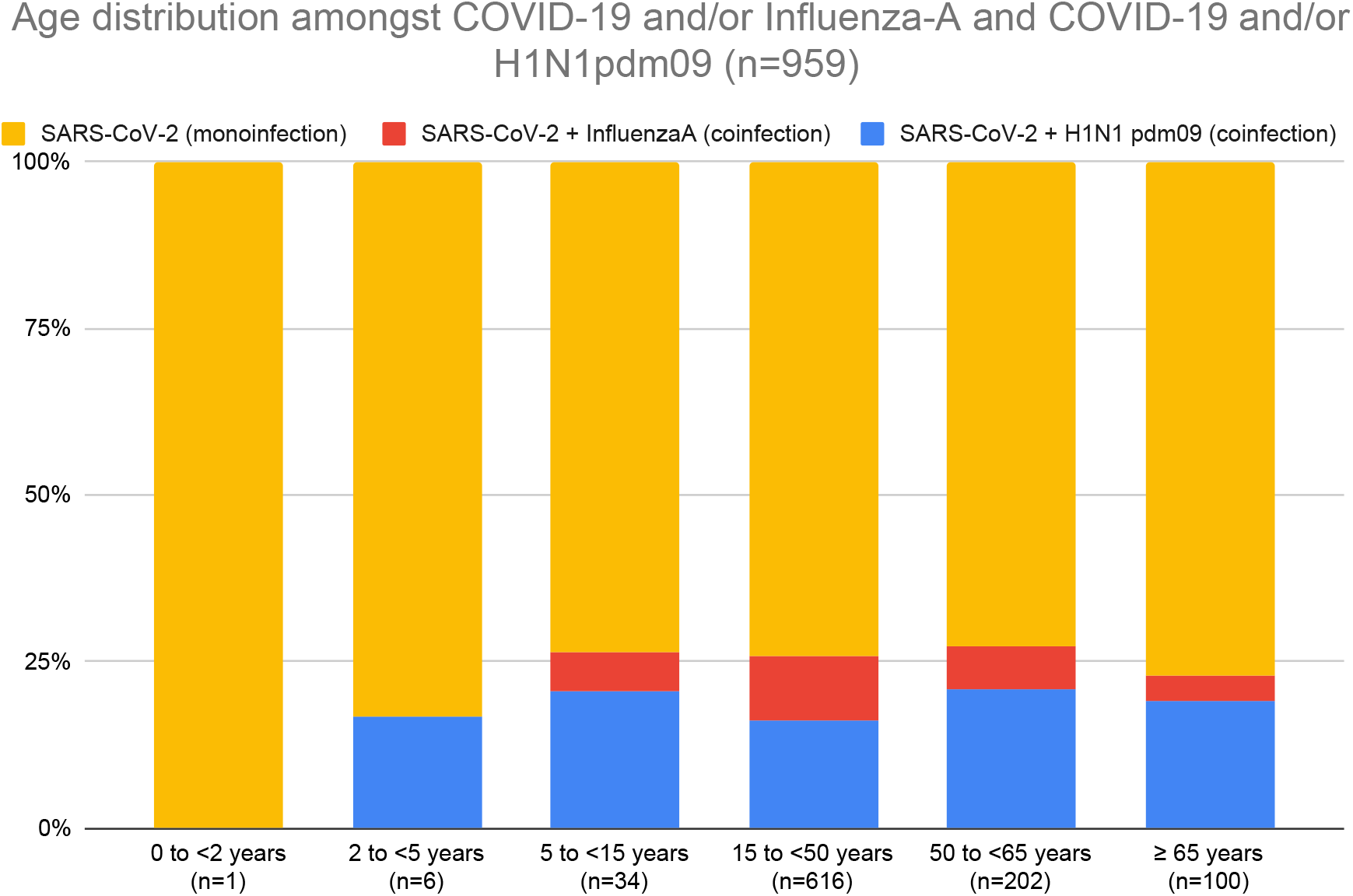
Percentage-wise distribution of SARS-CoV-2 and H1N1pdm09 coinfection (Blue); SARS-CoV-2 and Influenza A coinfection (Red); and SARS-CoV-2 mono-infection (Yellow) in different age groups of studied cases (n=959).

### 3.2 The comparative analysis of viral load among the coinfected cases

To understand the relative viral load in SARS-CoV-2 and Influenza A coinfection; and SARS-CoV-2 and H1N1pdm09 coinfection, a Ct values-based comparison among these coinfected samples were made. The Ct values of qRT-PCR correlate inversely with the viral load in the sample; a lower Ct value corresponds to a higher viral load and vice versa. The comparative analysis of Ct values showed that H1N1pdm09 consistently has a higher viral load in comparison to SARS-CoV-2 from May to December 2022 **(Fig.3a)** Furthermore, Influenza A consistently has a higher viral load in comparison to SARS-CoV-2 throughout the study period except during the month of August and October where the Influenza A viral load was lesser than that of SARS-CoV-2. **(Fig.3b)**. The comparative distribution of the Ct values of SARS-CoV-2, H1N1pdm09 and Influenza A in the tested samples was shown in **Figure 3c and Figure 3d**. In the majority of the SARS-CoV-2 and H1N1pdm09 coinfection cases, the Ct values of H1N1pdm09 throughout the study were observed to be low in comparison to SARS-CoV-2 Ct values. Similarly, in most of the SARS-CoV-2 and Influenza A coinfection cases, the Ct values of Influenza A throughout the study were observed to be low in comparison to SARS-CoV-2 Ct values.

**Figure 3a:**
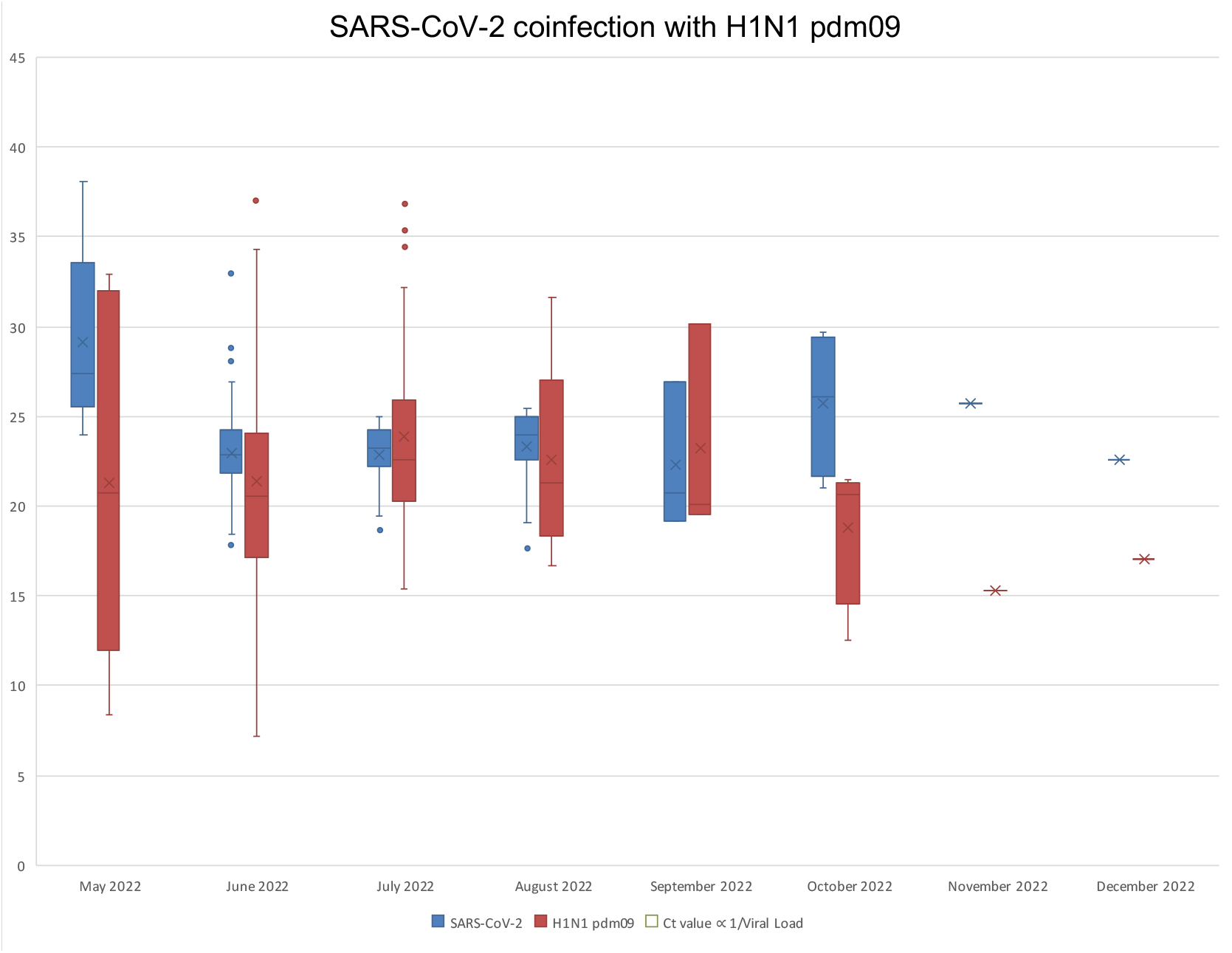
Monthwise viral load comparison between SARS-CoV-2 (Blue) and H1N1pdm09 (Red) in SARS-CoV-2 and H1N1pdm09 coinfection cases.

**Figure 3b:**
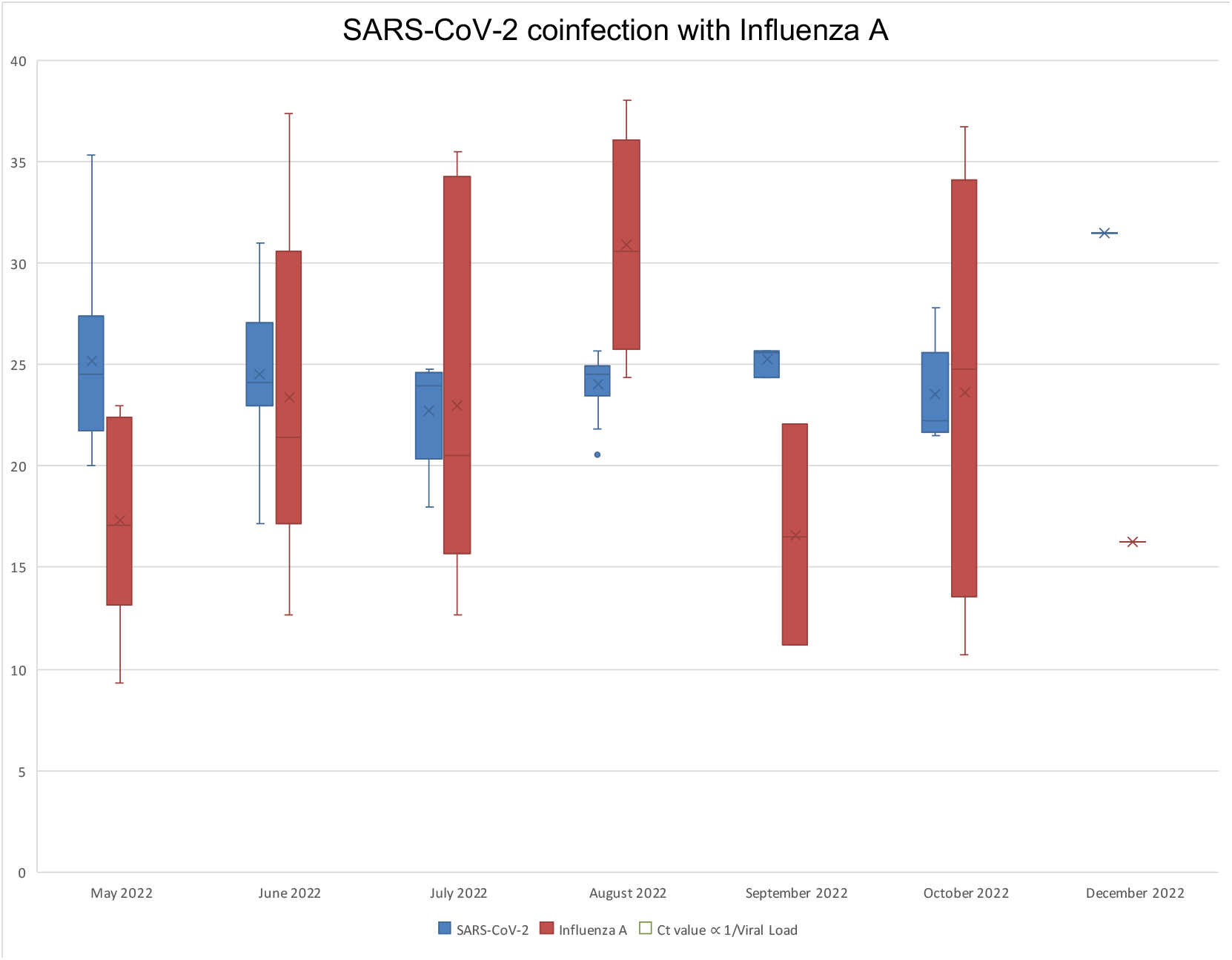
Monthwise viral load comparison between SARS-CoV-2 (Blue) and Influenza A (Red) in SARS-CoV-2 and Influenza A coinfection cases.

**Figure 3c:**
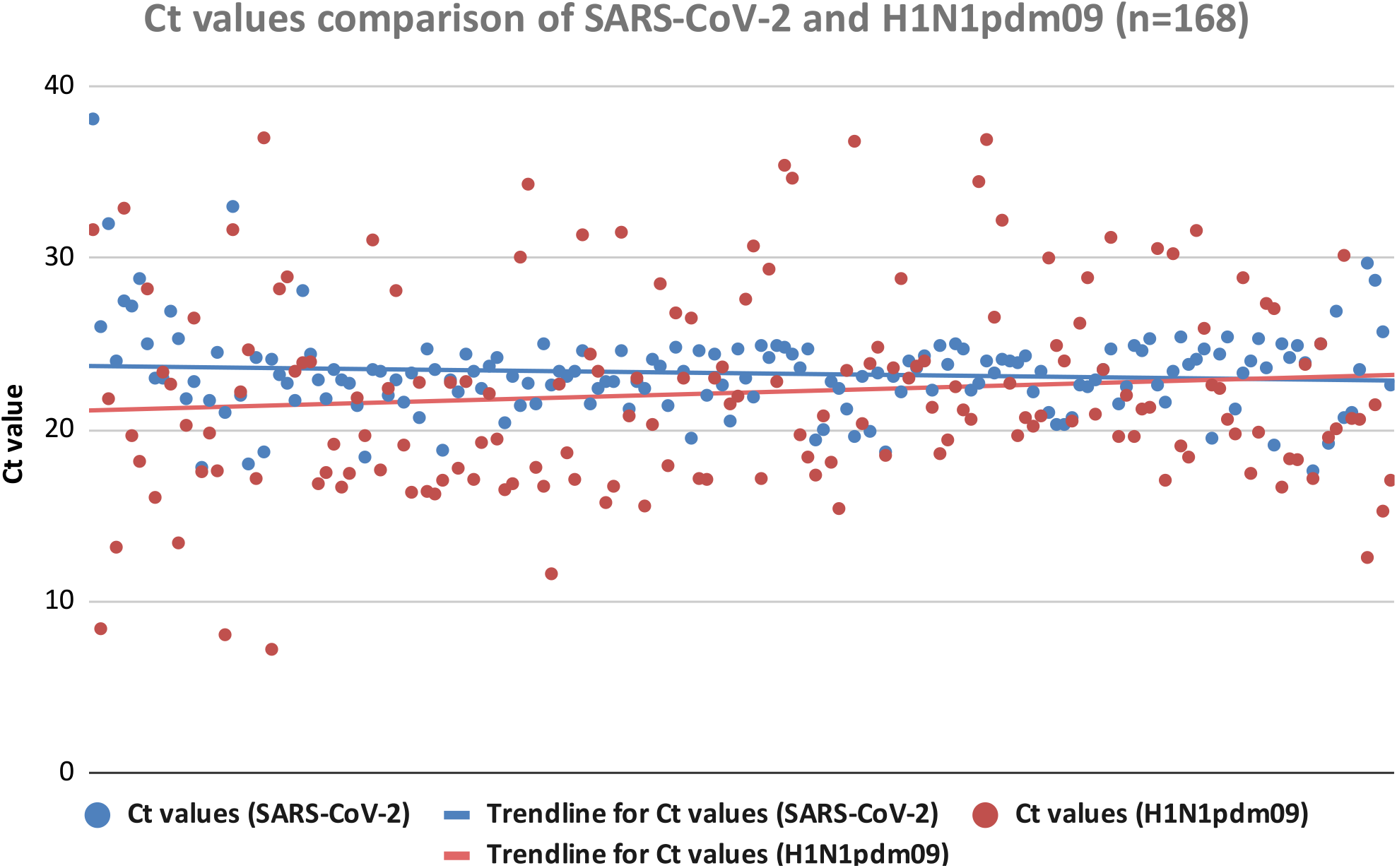
Distribution of Ct values of H1N1pdm09 (Red) and SARS-CoV-2 (Blue) for the study duration within SARS-CoV-2 and H1N1pdm09 coinfection cases (n=168).

**Figure 3d:**
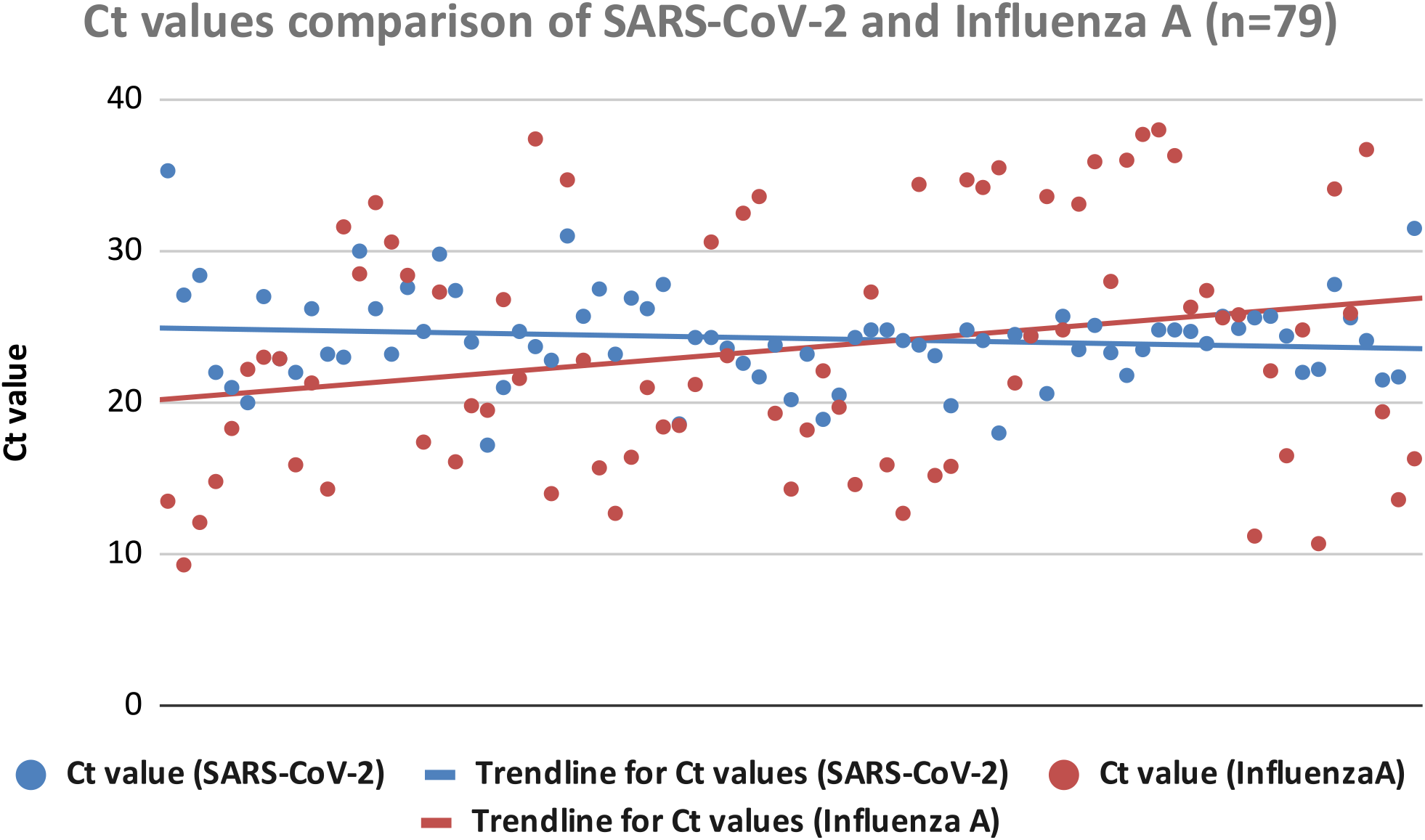
Distribution of Ct values of Influenza A (Red) and SARS-CoV-2 (Blue) for the study duration within SARS-CoV-2 and Influenza A coinfection cases (n=79).

### 3.3 SARS-CoV-2 variant distribution among the coinfected cases

Among the 959 SARS-CoV-2 samples, the WGS for SARS-CoV-2 was successfully performed for 815 samples, and the remaining 144 samples failed during the sequencing QC. In total, 46 types of SARS-CoV-2 variants were reported in the successfully sequenced samples, among these, the significant variants were BA.2.75 (27%), BA.2 (16%), BA.2.38 (10%), BA.2.12 (8%), BA.2.76 (5%), BA.5 (3%), BA.2.75.2 (3%), BA.2.74 (1%), BM.1.1 (1%), BA.2.75.1 (1%), B.1.1.529 (Omicron) (1%), and XBB (1%) (**Fig.4**). The major variants found among samples coinfected with Influenza A and SARS-CoV-2 (n=79) were BA.2 (27%), BA.2.75 (13%), BA.2.12 (11%), BA.2.38 (10%) and BY.1 (10%). The samples coinfected by SARS-CoV-2 and H1N1pdm09 (n=168) showed the significant variants BA.2.75 (29%), BA.2 (17%), BA.2.38 (13%), BA.2.12 (6%) and BA.5 (5%). The month-wise distribution of variants found in SARS-CoV-2 and Influenza A coinfection cases; and SARS-CoV-2 and H1N1pdm09 coinfection cases are shown in supplementary **figure S1**, and the percentage-wise distribution is shown in supplementary **figure S2**. The month-wise distribution of variants found in SARS-CoV-2 mono-infection cases is shown in supplementary **figure S3** and the percentage-wise distribution is shown in supplementary **figure S4**.

**Figure 4:**
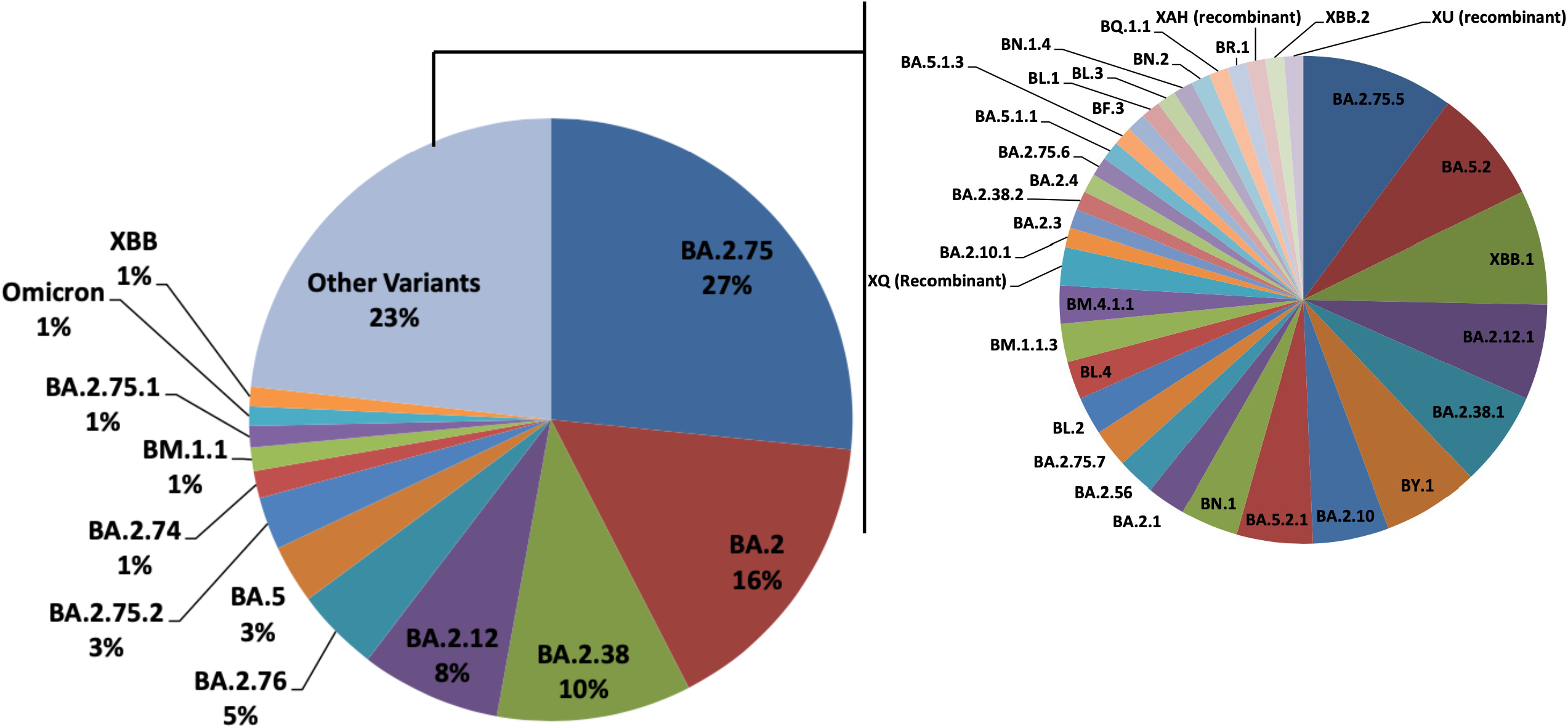
Overall SARS-CoV-2 variant distribution among the studied cases (n=959).

The SARS-CoV-2 variants identified by WGS in coinfection cases of SARS-CoV-2 and H1N1pdm09 collected from May to December 2022, revealed the significant variants viz BA.2, BA.2.38, BA.2.12, BA.2.74, BA.2.75, BA.2.76, BA.2.75.2, BA.2.75.5, BM.1.1, BY.1, BQ.1.1, XBB and XBB.1. The SARS-CoV-2 variants identified by WGS in coinfection cases of SARS-CoV-2 and Influenza A collected from May to December 2022, revealed the significant variants viz BY.1, BA.2.12, B.1.1.529, BA.2, BA.2.38, BA.2.74, BA.2.75, BA.2.76, BA.2.38.1, BA.5, BA.5.2, XU, BA.2.75.1, BM.1.1.3, BA.2.75.2, BA.2.10.1, BN.1.4, XBB and XBB.1. Furthermore, the SARS-CoV-2 variants identified by WGS in mono-infection cases of SARS-CoV-2 collected from May to December 2022, revealed the significant variants viz, B.1.1.529, BA.2.12, BA.2.38, BA.2, BA.5, BA.2.75, BA.2.76, BA.2.74, XAH, BA.2.75.2, BM.1.1, BA.2.75.1, XBB, XBB.1, BL.4, BN.1, XBB.2 and BA.5.2.1 (**Fig. 5**).

**Figure 5:**
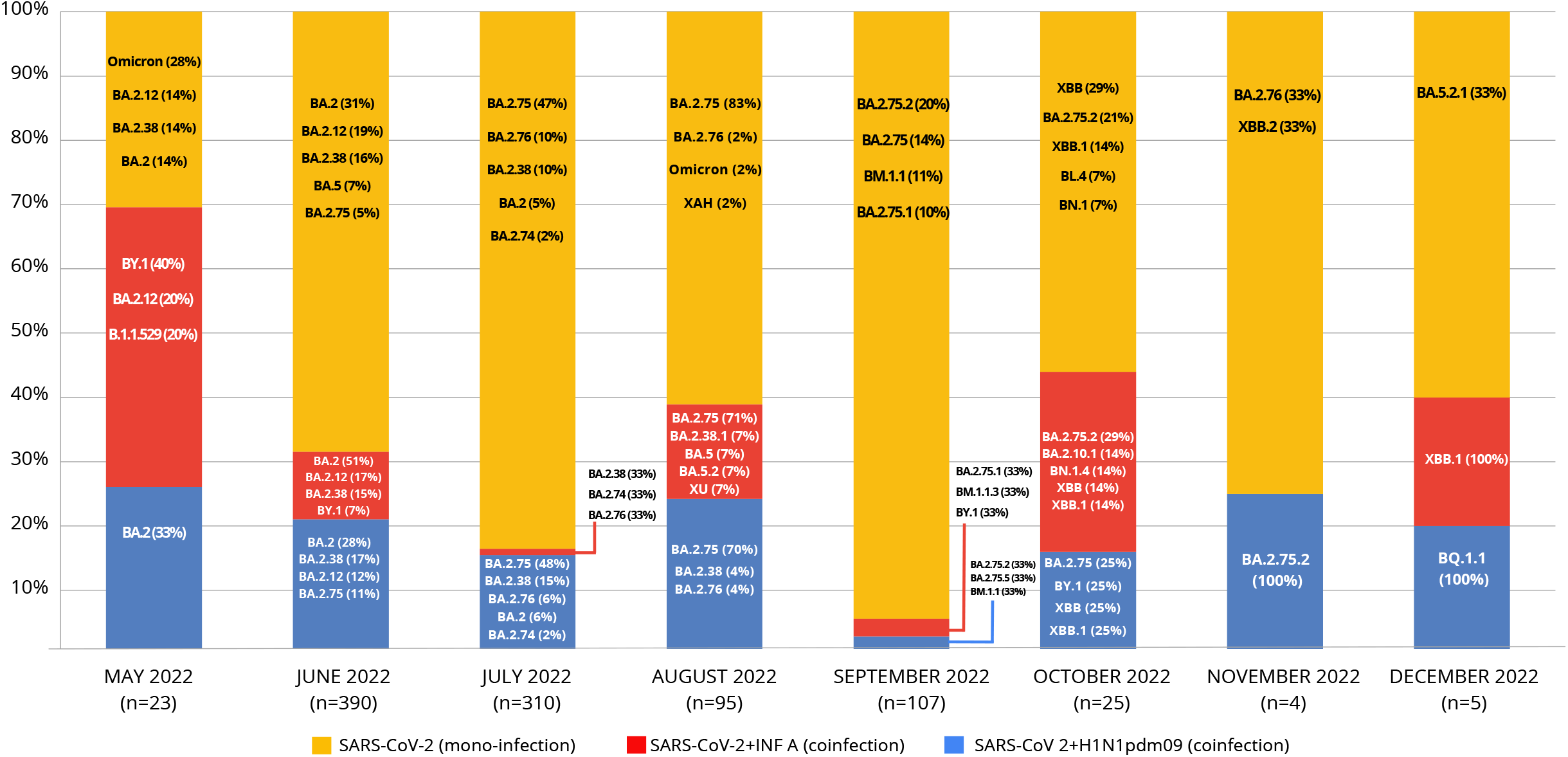
Monthwise percentage distribution of SARS-CoV-2 and H1N1pdm09 coinfection (Blue); SARS-CoV-2 and Influenza A coinfection (Red); and SARS-CoV-2 mono-infection (Yellow) cases from May 2022 to December 2022. The major SARS-CoV-2 variants identified by WGS are listed with the prevalence percentage.

### 3.4 Variant distribution among Symptomatic and Asymptomatic cases in the study group

Patient data of 959 SARS-CoV-2 positive samples showed that 514 patients were symptomatic and 445 were asymptomatic. In the SARS-CoV-2 and H1N1pdm09 coinfection group, 90 (54%) patients were reported symptomatic, and 78 (43%) were asymptomatic. The SARS-CoV-2 and Influenza A coinfection group showed 53 (67%) patients as symptomatic and 26 (33%) as asymptomatic. Symptomatic cases were found more in the SARS-CoV-2 and Influenza A coinfection group than in the SARS-CoV-2 and H1N1pdm09 coinfection group. The major SARS-CoV-2 variants identified by WGS among the symptomatic and asymptomatic cases in the SARS-CoV-2 and H1N1pdm09 coinfection group, and SARS-CoV-2 and Influenza A coinfection group are summarised in **Figure 6**. The major variants found in the SARS-CoV-2 and H1N1pdm09 coinfection group for both symptomatic and asymptomatic cases were observed to be similar. The majority of the symptomatic and asymptomatic cases in the SARS-CoV-2 and Influenza A coinfection group also showed a similar variant distribution except BA.5 in symptomatic cases and BA.2.38.1 in asymptomatic cases.

**Figure 6:**
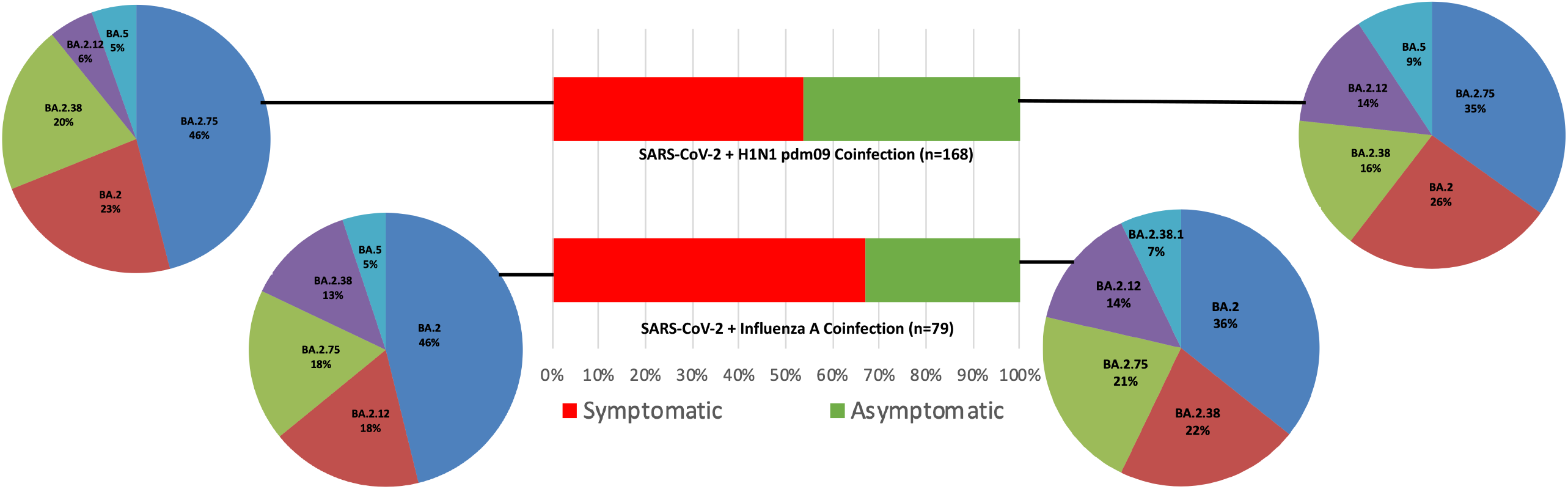
Distribution of the significant SARS-CoV-2 variants identified by WGS within symptomatic and asymptomatic cases of SARS-CoV-2 and H1N1pdm09 coinfection, and SARS-CoV-2 and Influenza A coinfection.

## 4. DISCUSSION

This study revealed a significant number of coinfection with H1N1pdm09 and Influenza A (other subtypes) in the SARS-CoV-2 positive samples collected for genomic surveillance in India during the period of May to December 2022. This period also resonates with the peak of the H1N1pdm09 cases in 2022. This study revealed that during this timeline, out of the total SARS-CoV-2 positive samples, 26% of the samples had coinfection with Influenza A (including 18% H1N1pdm09, a subtype of Influenza A and 8% of Influenza A other subtypes). In this study, the H1N1pdm09 and SARS-CoV-2 coinfection cases were significantly found during the period May to December 2022, this observation resonates with the prevalence of H1N1pdm09 reported by ICMR during the same period in India [15].

When the coinfected cases were analysed as per the age distribution, a high number of H1N1pdm09 cases was found in the age group of 15 to <50 years compared to the other age groups. A meta-analysis of SARS-CoV-2 and Influenza coinfections studies by Dadashi et al. 2021 showed that most of the coinfections were from the age group of >50 years, followed by the age group <50 years [24]. It is essential to consider that the death rate and birth rate are considerably higher in India, leading to a significantly large population in the middle-aged group, which leads to maximum coinfection cases falling within the age group of 15 to 50 years. However, the proportion of coinfection cases remains highest in 50 to <65 years. There is a linear correlation between the comorbidities and increasing age, such observations explain a higher vulnerability to infections in the ageing population.

The comparative analysis of Ct values among the coinfection cases revealed that the viral load of H1N1pdm09 was higher in comparison to the viral load of SARS-CoV-2. Similarly, the viral load of Influenza A was higher as compared to the viral load of SARS-CoV-2. The viral load estimation in the samples can give an insight into the etiological agent contributing to the severity of the disease as well the transmission dynamics of the viruses. The higher viral load of the H1N1pdm09 and Influenza A in the coinfected cases may be the underlying reason for the symptomatic clinical presentation in these patients. Additionally, the higher viral load of the H1N1pdm09 and Influenza A in the SARS-CoV-2 coinfected cases could also be due to existing cohort immunity against SARS-CoV-2 due to SARS-CoV-2 vaccination and/or natural immunity against SARS-CoV-2 in the Indian population.

When the COVID-19 pandemic began in December 2019, it started the pandemic with one variant of SARS-CoV-2 known as the Wuhan-Hu-1 strain. As the COVID-19 pandemic progressed, the SARS-CoV-2 virus mutated into several variants like Alpha, Beta, Gamma, Delta and Omicron. It is possible to identify the circulating SARS-CoV-2 variants during the pandemic only by the SARS-CoV-2 WGS approach, which is the existing gold standard. The SARS-CoV-2 WGS results showed that all 46 types of the variants detected were sub-lineages of Omicron, which is associated with asymptomatic or mild symptoms in SARS-CoV-2 infections. Therefore, the findings of the SARS-CoV-2 WGS further reaffirm that amongst the coinfection cases, the H1N1pdm09 and Influenza A viruses could be more contributing in terms of viral pathogenesis and the resultant clinical manifestations.

When the symptomatic and asymptomatic cases were compared, Influenza A and SARS-CoV-2 coinfection groups showed relatively higher symptomatic cases than H1N1pdm09 and SARS-CoV-2 coinfection groups. Furthermore, the distribution of significant SARS-CoV-2 variants among the symptomatic and asymptomatic cases in the coinfected cases was also similar. Although the qRT-PCR quantitatively confirmed the relative viral loads of SARS-CoV-2, Influenza A and H1N1pdm09 in the positive samples, the SARS-CoV-2 WGS allowed us to resolve the type of SARS-CoV-2 variant present in these samples to be of omicron sublineage. This allowed us to infer that the clinical manifestations of these coinfected cases may be primarily due to H1N1pdm09 and Influenza A viral infections. SARS-CoV-2, Influenza A and H1N1pdm09 are respiratory pathogens often exhibiting similar etiological characteristics in infected patients. Additionally, as these viruses show similar pathogenesis and symptoms of ILI/SARI in the patients, it can be difficult to distinguish based on clinical manifestation and without proper deployment of the diagnostic tests. Furthermore, as these pathogens target the respiratory system, infection with either of these viruses can make the patient more prone to infection with the other virus [25]. Co-infections with these viruses can also aggravate symptoms and cause complications in the patient [11, 25]. Therefore it is essential to understand the prevalence of such co-infections in large populations for proper mitigation strategies.

## 4. CONCLUSION

This study brings forth the relevance of coinfection analysis for the H1N1pdm09 virus and Influenza A virus alongside SARS-CoV-2 while investigating the etiological agent amongst the patients manifesting ILI/SARI symptoms. As this study has been performed retrospectively on the already confirmed and reported cases of SARS-CoV-2, it is essential to note that almost one-fourth of the total SARS-CoV-2 reported cases had been found to be coinfected with H1N1pdm09 or Influenza A. Notably, the overall viral load of the H1N1pdm09 or Influenza A had been more when compared to the SARS-CoV-2 in the coinfected cases, implying that H1N1pdm09 or Influenza A being the main contributing factor towards the clinical manifestation of ILI/SARI. The aforesaid observation was further supported by the finding that no significant correlation of the SARS-CoV-2 variants identified by WGS in the coinfection cases could be established with clinical symptoms as all the SARS-CoV-2 variants belonged to the Omicon lineage, which is most commonly associated with the asymptomatic or mild symptoms of SARS-CoV-2. We believe that the high sensitisation towards SARS-CoV-2 during the COVID-19 pandemic has caused a bias leading to underplaying the other etiological agents contributing to the disease outcome. Also, it is worthwhile to note that there is an existing cohort immunity against SARS-CoV-2 amongst the existing population due to the vaccination and natural infection, similar cohort immunity may not exist against Influenza A virus in the given population, and this may lead to higher incidences of Influenza A infections in the given population. It is proposed that while ascertaining the aetiology of ILI/SARI, the investigation for H1N1pdm09 and Influenza A also be carried out simultaneously by using diagnostic methods like qRT-PCR to understand the dynamics of the viral load and variant identification by deploying NGS methods for arriving at a logical interpretation, especially in the cases of coinfection for taking an informed decision during the patient management and treatment discourse.

## Data Availability

All data produced in the present study are available upon request

## FIGURE LEGENDS

Figure S1: Monthwise distribution of SARS-CoV-2 variants identified by WGS in coinfection cases of SARS-CoV-2 and H1N1pdm09 (n=168) and coinfection cases of SARS-CoV-2 and Influenza A (n=79).

Figure S2: Monthwise percentage distribution of SARS-CoV-2 variants identified by WGS in coinfection cases of SARS-CoV-2 and H1N1pdm09 (n=168) and coinfection cases of SARS-CoV-2 and Influenza A (n=79).

Figure S3: Monthwise distribution of SARS-CoV-2 variants identified by WGS in mono-infection cases of SARS-CoV-2 (n=712).

Figure S4: Monthwise percentage distribution of SARS-CoV-2 variants identified by WGS in mono-infection cases of SARS-CoV-2 (n=712).

## CONFLICT OF INTERESTS

The authors declare no conflicts of interest.

## ACKNOWLEDGMENTS

This work was supported and funded by CSIR-NEERI, Nagpur. It is certified that the manuscript has been checked for plagiarism by the institute knowledge resource centre through iThenticate (anti-plagiarism software) KRC No. CSIR-NEERI/KRC/2023/FEB/EVC/1

## Notes

### Competing Interest Statement

The authors have declared no competing interest.

### Funding Statement

This study was funded and supported by the Council of Scientific and Industrial Research - National Environmental Engineering Research Institute, Nagpur

### Author Declarations

Council of Scientific and Industrial Research-National Environmental Engineering Research Institute (CSIR-NEERI), Nagpur, India. Institutional Ethics Committee (IEC)

