## Supplementary figures and images for "Dynamics of Influenza A and SARS-CoV-2 coinfections during the COVID-19 pandemic in India"

### Supplementary Figure 1

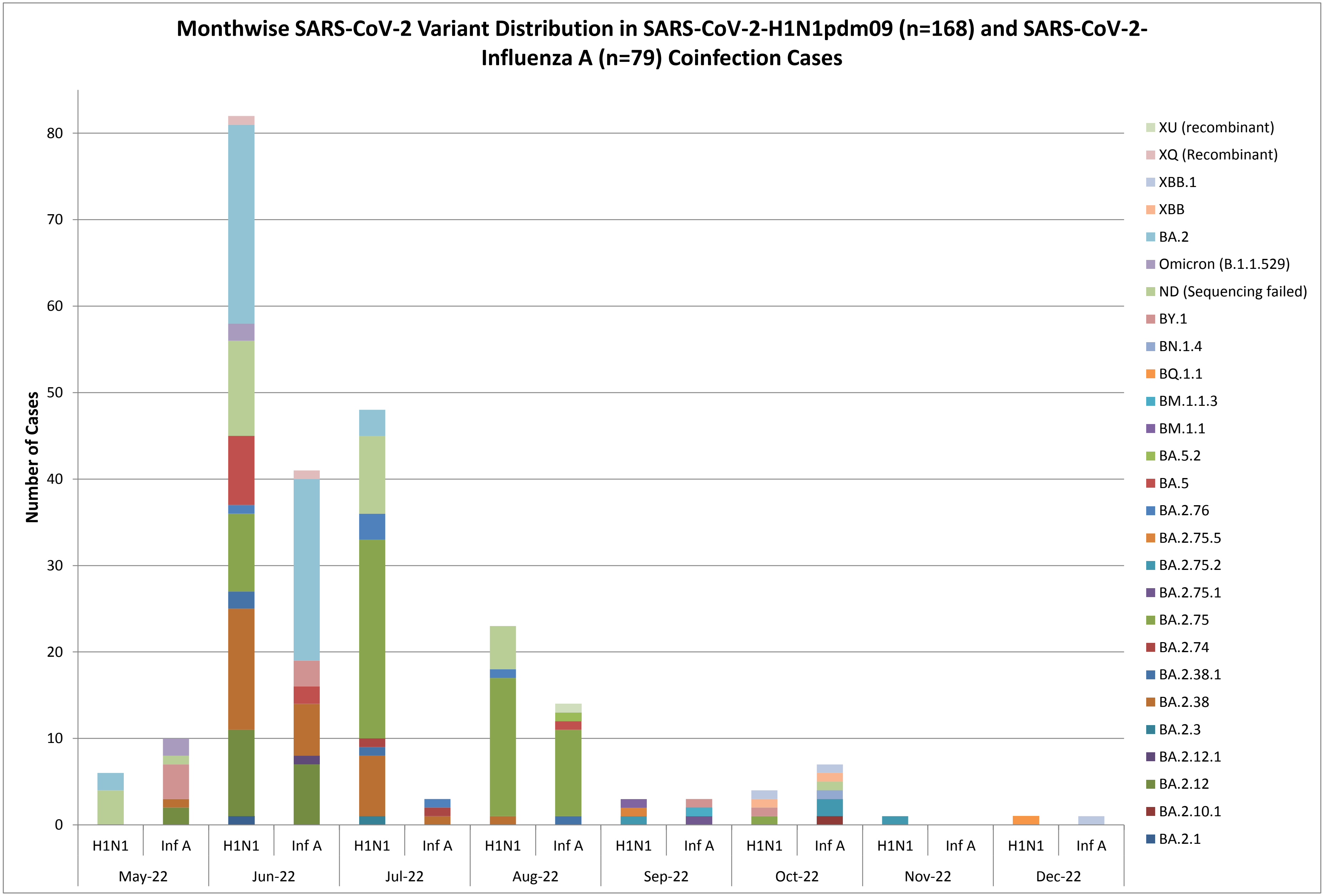

### Supplementary Figure 2

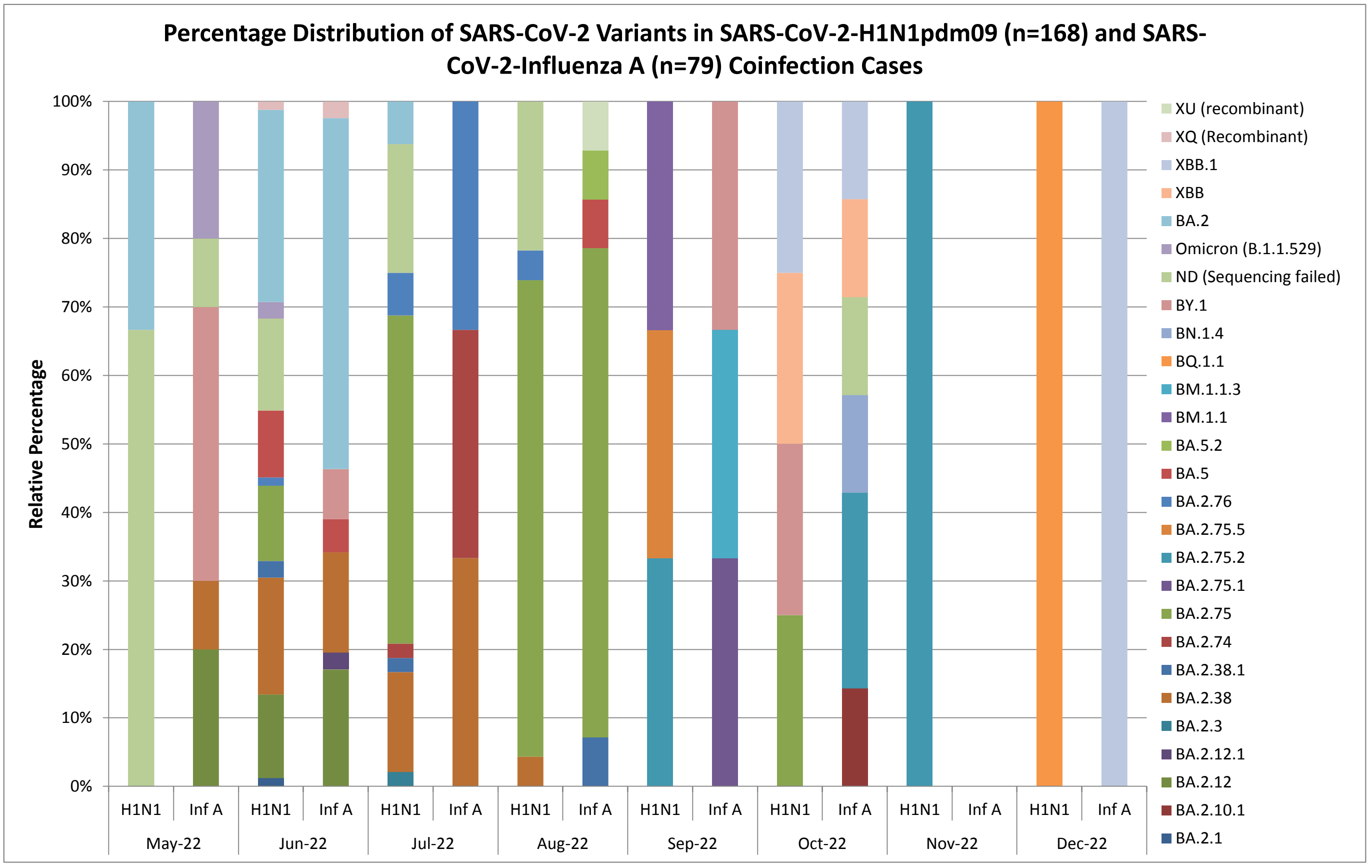

### Supplementary Figure 3

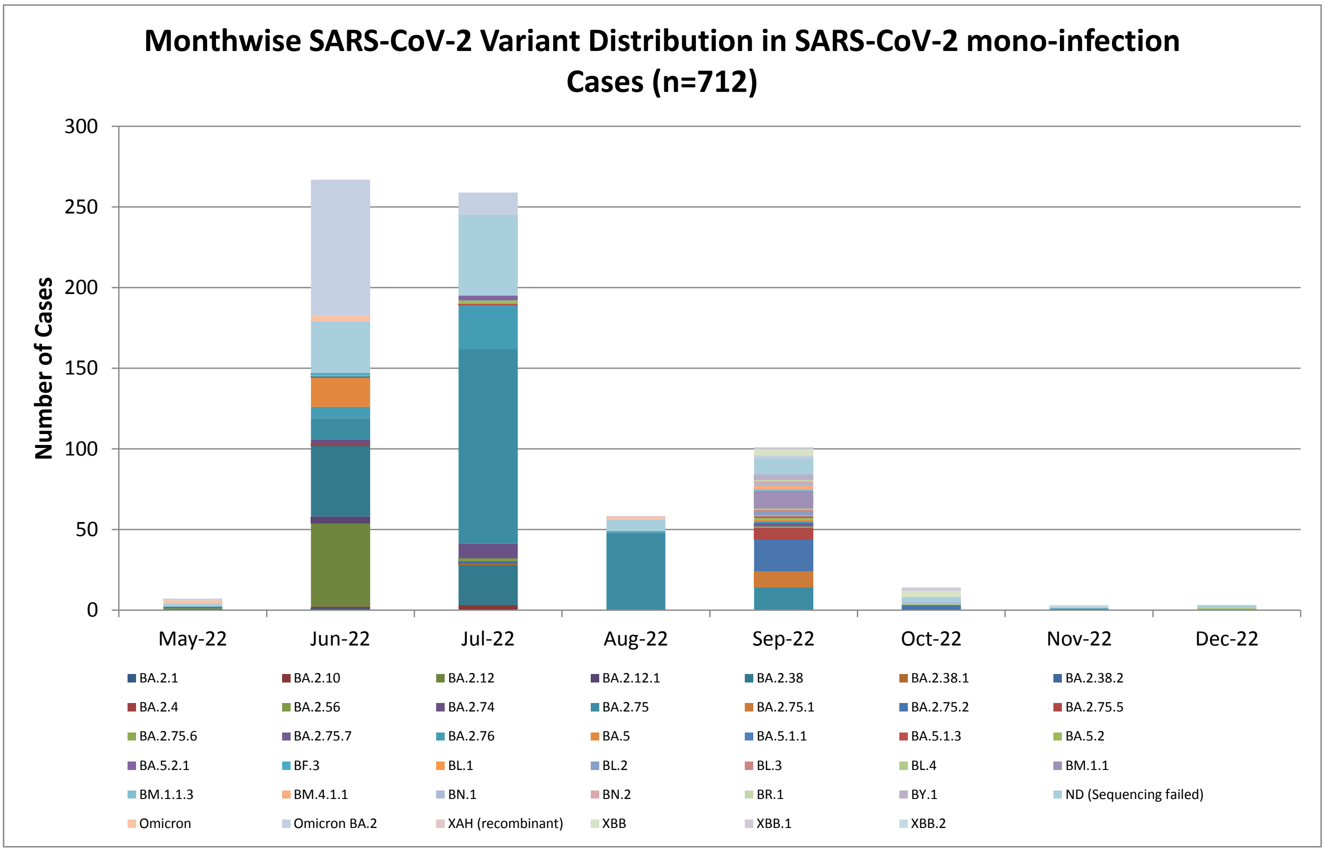

### Supplementary Figure 4

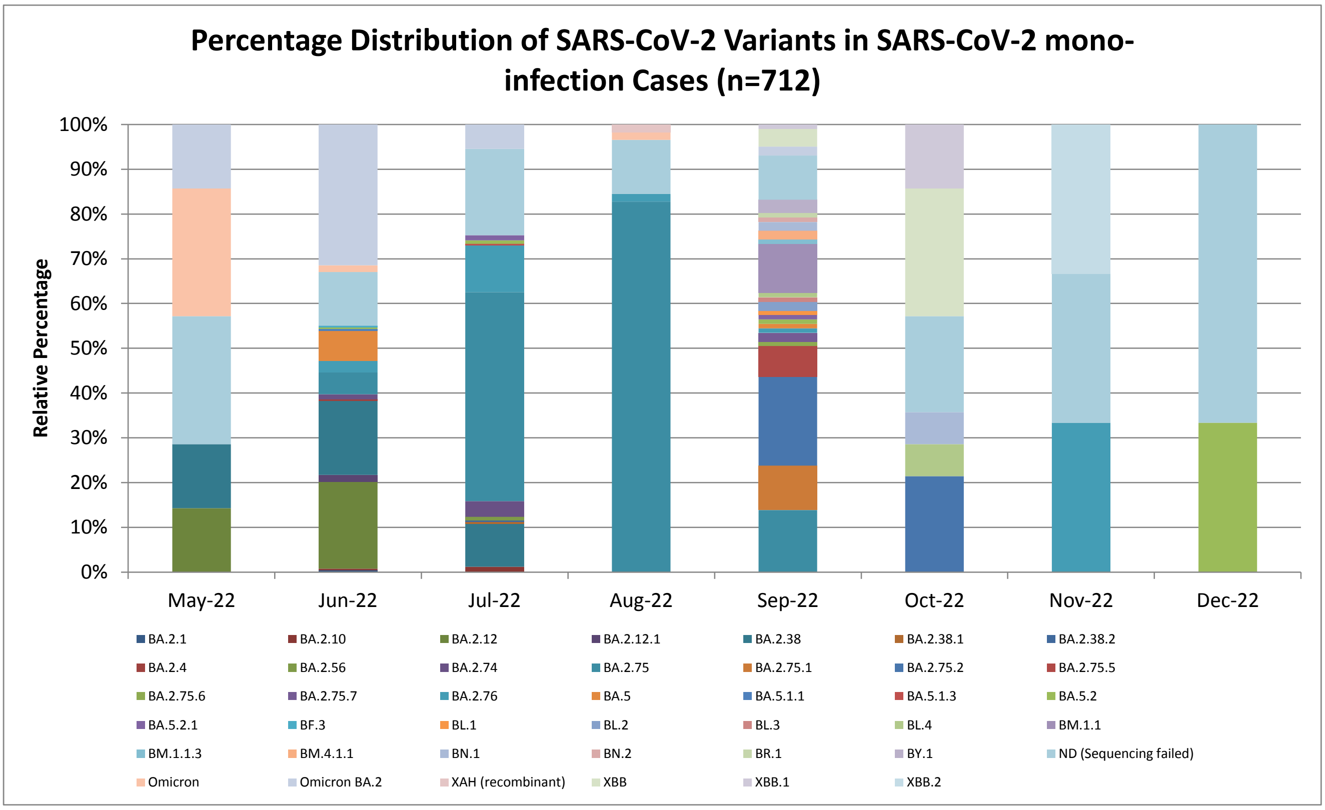
